# Covid 19 and orthopaedic surgery in a large trauma centre in India

**DOI:** 10.1101/2020.09.05.20188920

**Authors:** Kannan Karuppiah Kumar, M N Kumar, Muniramaiah Ravishankar, Thomas Chandy, Chetan Rai, A Chetan, Vijay Girish, Krishna Kumar, Praveen S Battepati, Deepak G Shivarathre, Harish Puranik, Noel Naleen Kumar, Krishan Prasad, Dr Harshvardhan, N K Deepu, Mayur Shetty

## Abstract

**Background:** We are in the midst of a pandemic caused by the novel SARS-Cov-2 virus. A large percentage of the patients are asymptomatic and hospitals around the world are struggling to restart routine services. We report the results of a universal testing protocol of all patients who underwent orthopaedic surgery in the month of July 2020 in a large orthopaedic speciality hospital in Bangalore, India.

**Methods:** A retrospective study of all patients who underwent orthopaedic surgery in the month of July 2020 at a tertiary care orthopaedic speciality hospital in Bangalore, India. All patients underwent nasopharyngeal swab test before surgery. A questionnaire was used to assess the patient before the RT-PCR nasopharyngeal swab test. Data regarding imaging, investigations and follow up was recorded.

**Results:** In the month of July 2020, 168 patients underwent routine nasopharyngeal RT-PCR swab test for COVID-19 prior to planned orthopaedic surgical procedure (Both trauma and elective cases). 16 of the RT-PCR tests were positive. However vascular cases and absolute emergencies were done without a RT - PCR test with PPE and all universal precautions. 11 patients underwent emergency surgery without a RT-PCR test. All 16 cases who were positive were asymptomatic. The asymptomatic positive rate was 9.52%. Of the 11 patients who underwent emergency surgery without a RT-PCR test, only one patient had a positive test post-operatively.

**Conclusions:** Routine nasopharyngeal RT-PCR testing revealed a high rate of asymptomatic cases. If the RT-PCR test is positive, it is best to defer the case till the test returns negative. All precautions must be taken while performing emergency surgeries. Our algorithm in managing patients has proven to be effective and can be replicated with ease to continue operating and taking care of orthopaedic patients during this pandemic.

## Introduction

We are in the midst of a pandemic. A lot of routine orthopaedic work has come down, and hospitals have been finding it difficult to restart work. Covid-19 is raging through Bangalore and India for the last few months. We are a large tertiary care hospital specialized in orthopaedic surgery. We have modified our protocols and routinely obtained a RT-PCR for all non emergency cases before surgery. If there was an absolute emergency we went ahead with the procedure without performing RT-PCR after taking adequate precautions including full PPE protection. We also made sure that we reduced the operating time to as little as possible. We present our algorithm for managing routine orthopaedic patients.

## Methods and materials

We obtained informed consent from all patients. All patients who underwent RT-PCR before non-emergent (trauma) or elective surgery in the month of July 2020 were included in the study. This information was obtained from electronic records available at our hospital. We also recorded any information available regarding symptoms (Cough, cold, fever, sore throat, breathlessness and anosmia) before obtaining the swab. All positive patients were followed up till they turned negative.

Management algorithm (Figure 1) – All patients coming to the hospital and had an indication for surgery were triaged based on the emergency(Table 1)(1). Based on the triage only absolute emergency cases were included for surgery (11 cases). If it was an absolute emergency, the patient was taken up for surgery in an isolation operation theatre with all PPE precautions. Once the surgery was done, a nasopharyngeal swab was sent at the earliest for RT-PCR. If the case is triaged as a non emergency, the patient is admitted to the isolation ward after appropriate first aid and splinting. Till the time that the report of RT-PCR is received, the patient is assumed to be Covid - Positive. Once the RT-PCR report has been received and is negative, the patient is taken up for surgery, again with adequate precautions.

**Table 1:** What is an absolute emergency?- where surgeries can be done without RT-PCR

|  |  |
| --- | --- |
| Trauma | 1. Severe crush injury with an unstable patient<br>2. Vascular injury<br>3. Infection/Septic arthritis |
| Pediatric | 1. Widely displaced supracondylar fractures<br>2. Severe pain<br>3. Vascular injuries |
| Hand | 1. Replants and revascularizations |
|  | 2. Crush injuries |
| Spine | Cauda equina<br>Trauma with fracture instability |

**Figure 1:**
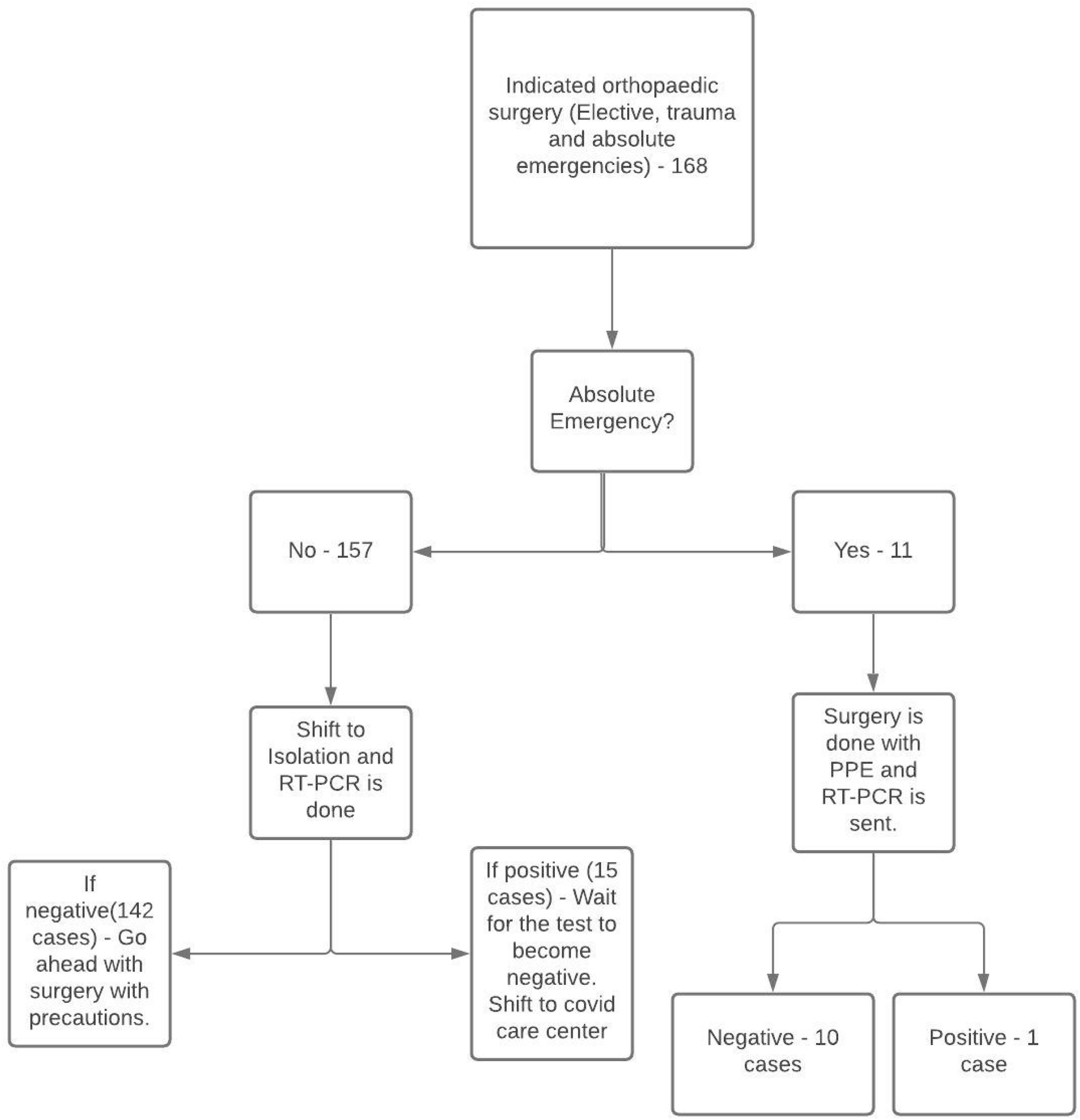
Algorithm of how orthopaedic cases were managed in the month of July 2020.

If the RT-PCR is positive, the patient is transferred to a covid care center or home. Once the patient is RT-PCR negative, the elective/non emergency surgery is performed.

## Results

168 patients presented to our hospital for surgical care (elective and trauma patients). 112 were male and 46 female. 11 of these patients underwent emergency surgery without RT-PCR results (6.5% of the cases). All of these 11 patients were asymptomatic and had their swabs taken after the emergency surgery(Table 2). One out of these 11 cases turned out to be positive. This was a suspected case of vascular injury to the thumb, but underwent K wiring and debridement. He was asymptomatic and was discharged for appropriate Covid care. He needed a small flap cover for the thumb and this was done after he became RT-PCR negative. The other 10 patients were RT-PCR negative post operatively.

**Table 2:**
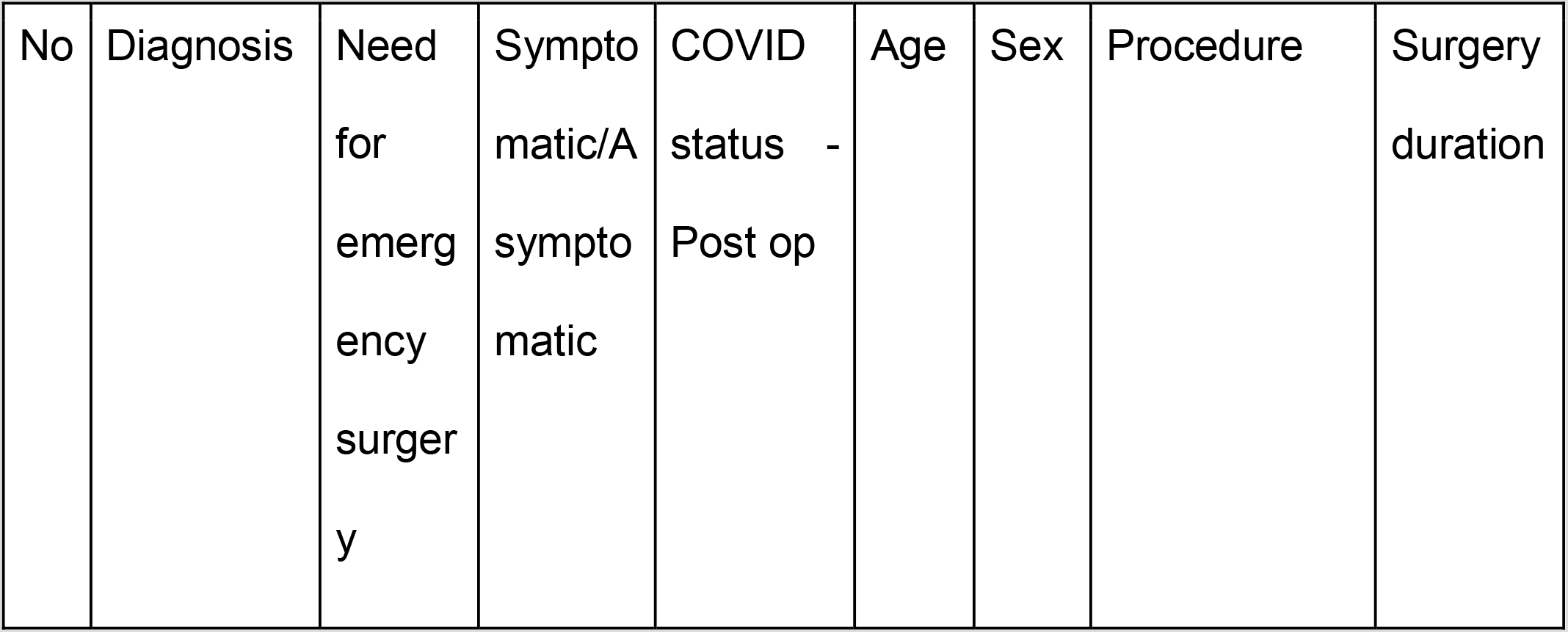
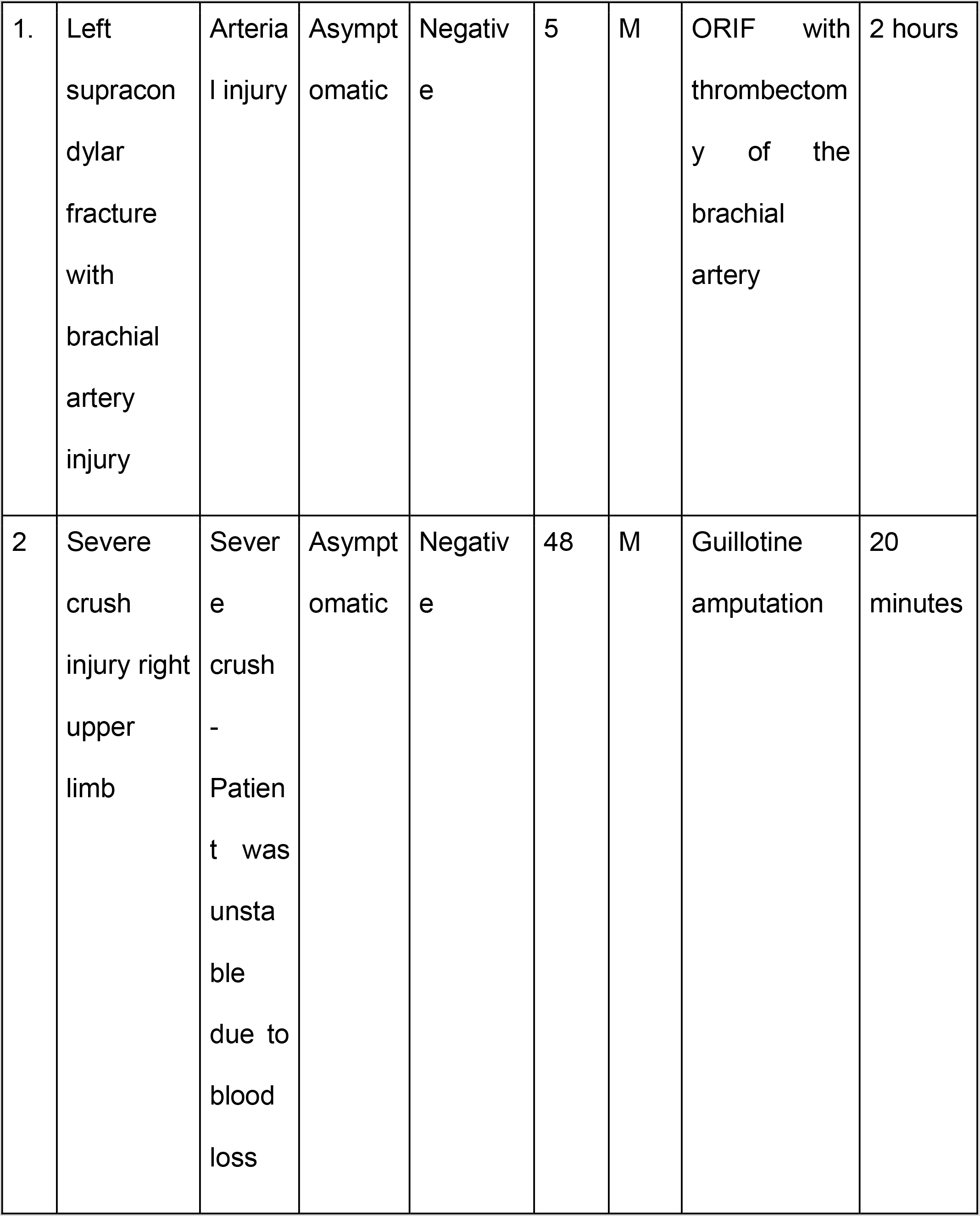
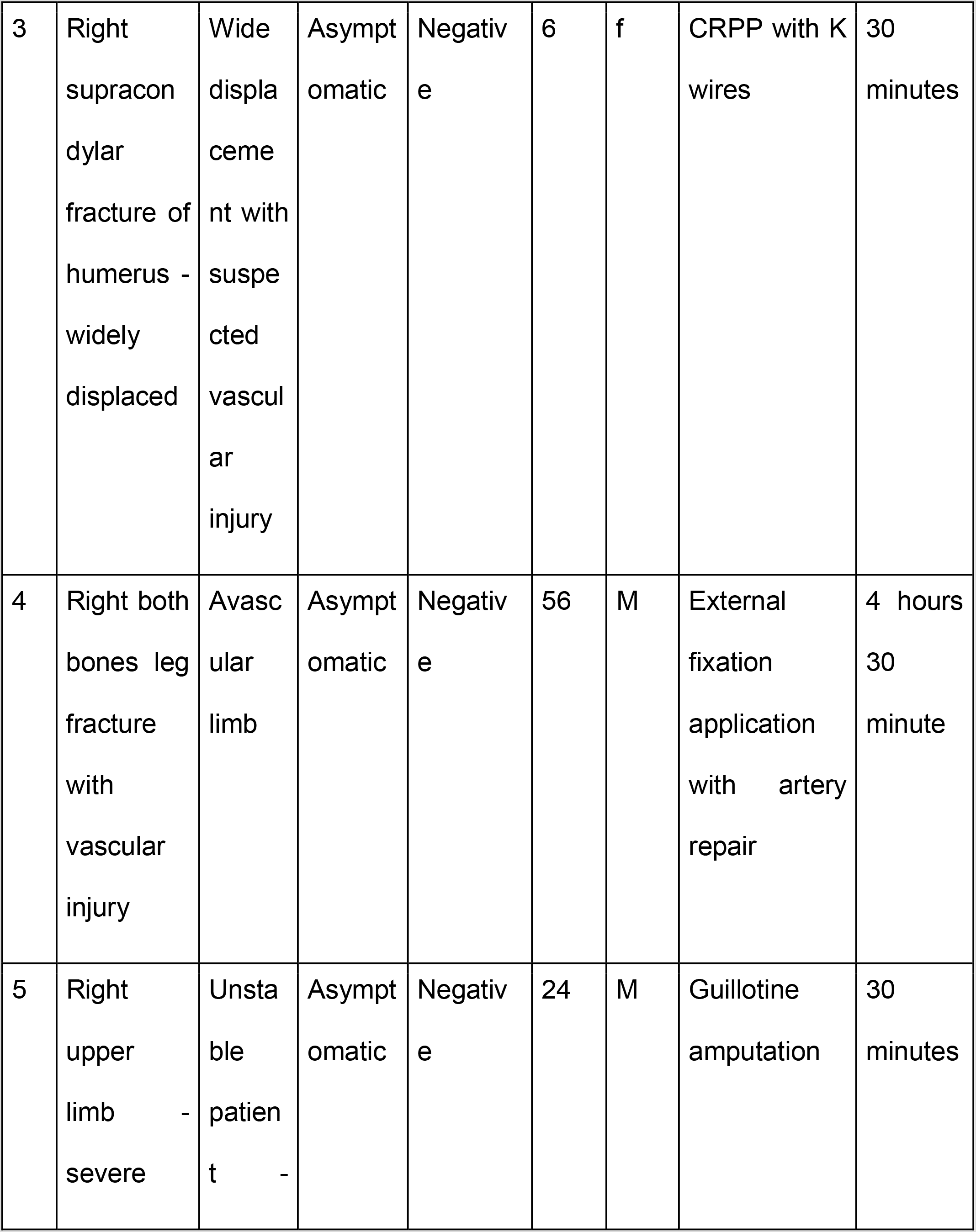
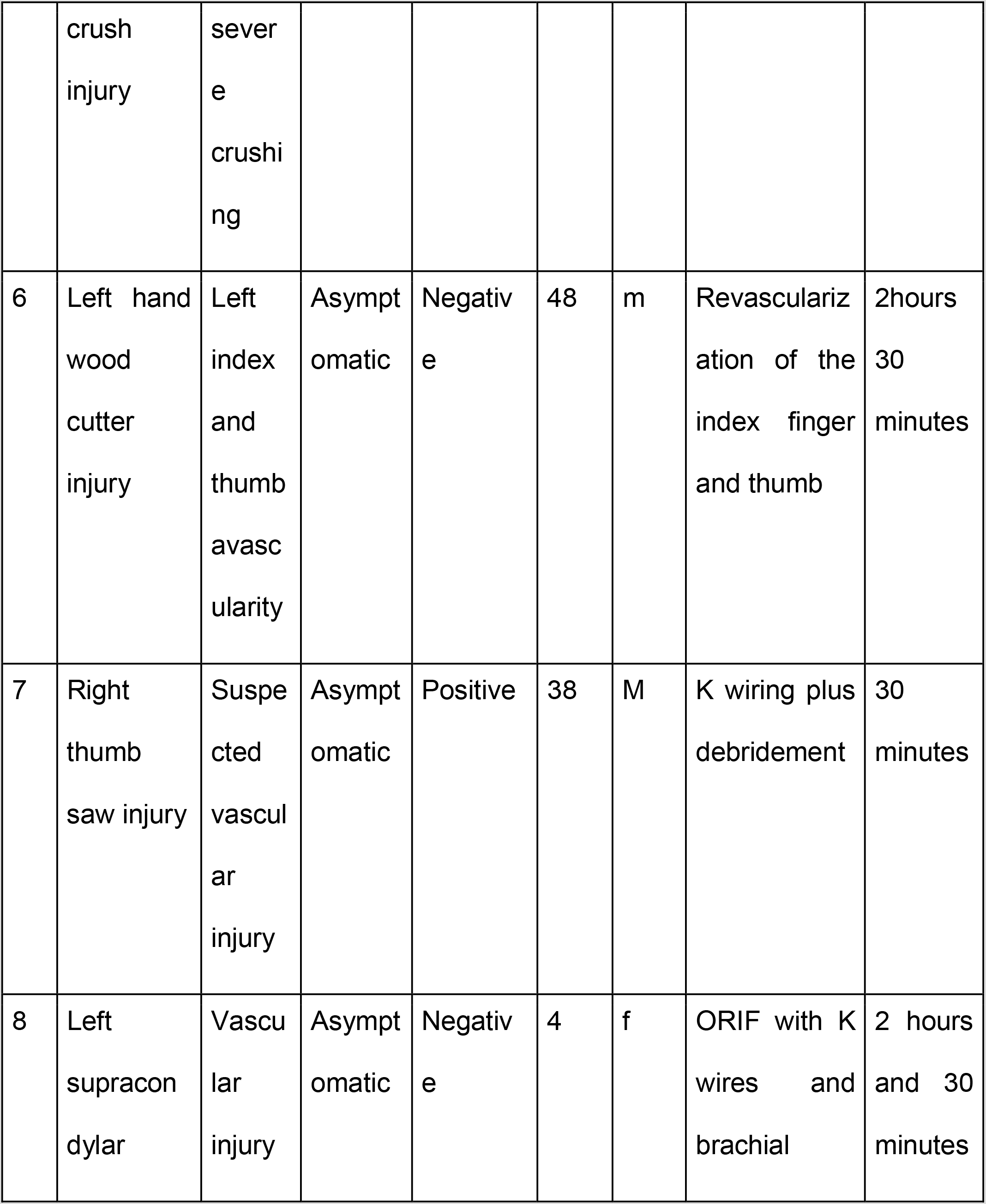
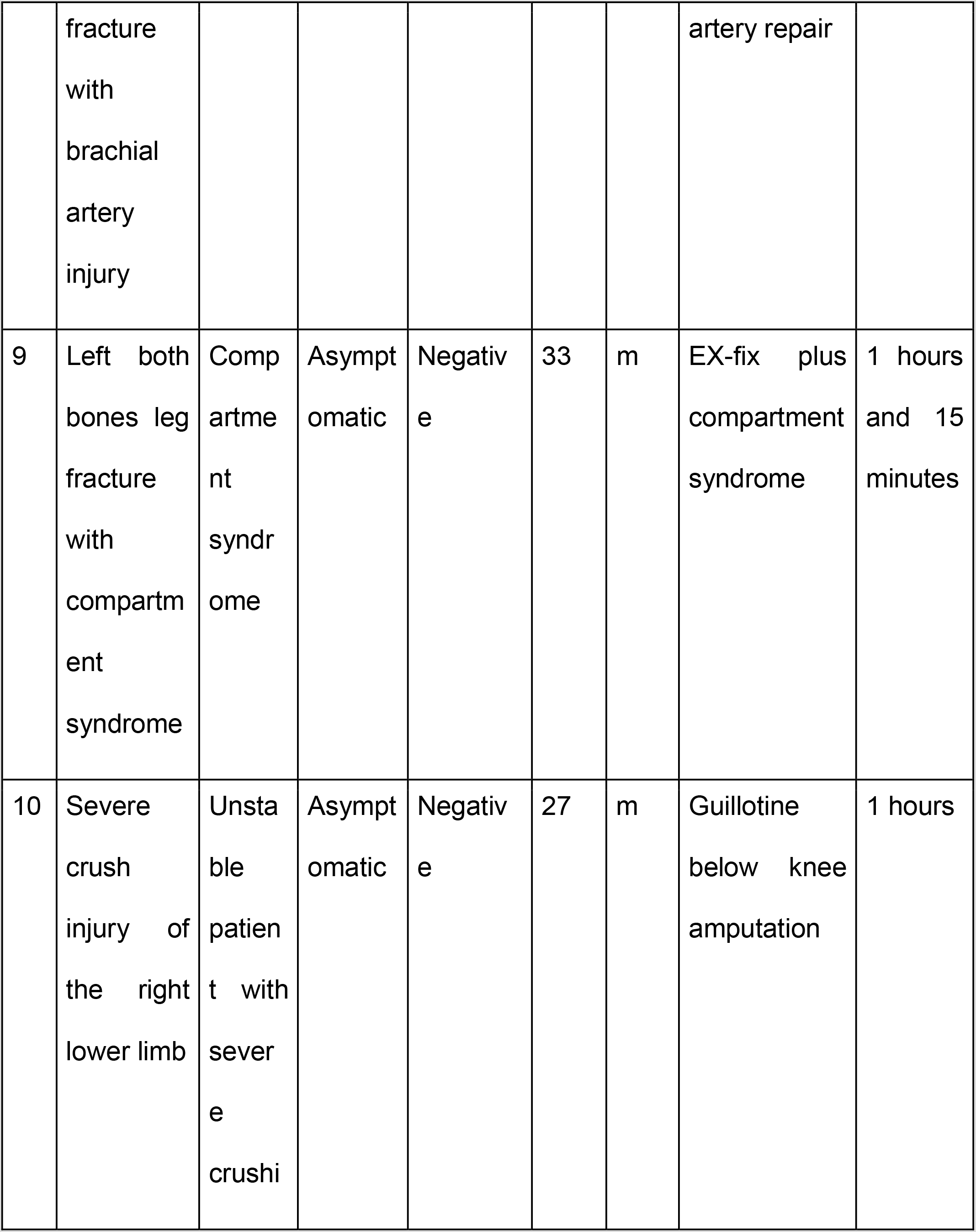
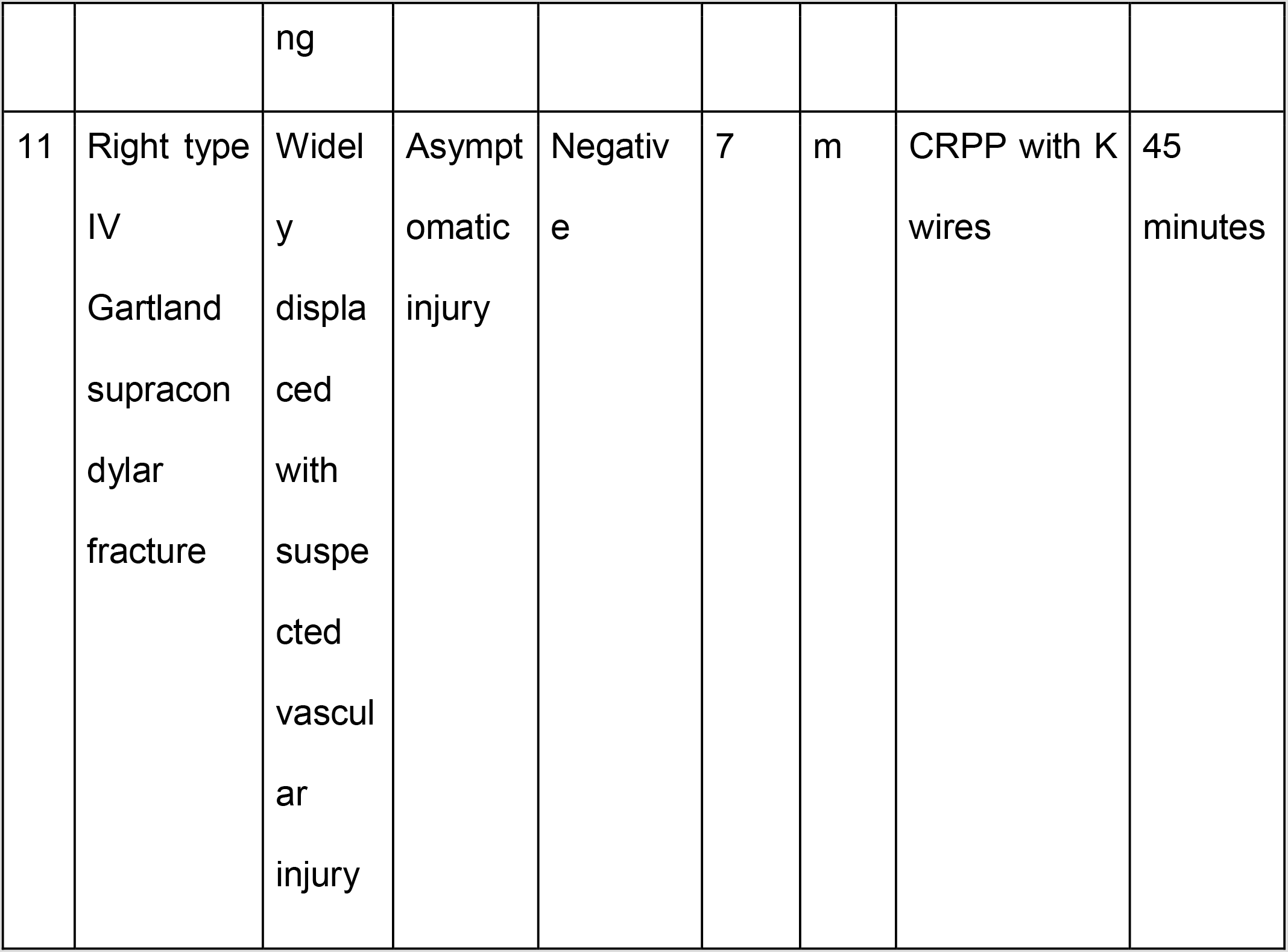
Emergency cases done without RT-PCR test

Out of the rest of the 157 patients who underwent RT - PCR testing before surgery. The results of 15 patients came back positive. One patient (both bones forearm fracture) was lost to follow up. Three patients (One IT fracture, one distal end radius fracture and one forearm fracture) opted to not have surgery after the RT-PCR came back positive. 11 patients had surgery once the RT-PCR became negative. The average time spent in doing an emergency case was 90 minutes (range 20 minutes - 4hours 30 minutes). All chest Radiographs were normal.

We performed a total of 160 surgeries in the month of July 2020. The asymptomatic positive rate in our center was 9.52% (i.e 16 cases out of 168) (July 2020, Bangalore).

Only one patient was RT-PCR positive at the time of surgery. This surgery was done in a dedicated OR and all personnel were using PPE. The surgeon, assistant surgeon and scrub nurse were quarantined for 7 days (According to protocol) after this surgery.

## Discussion

We have used nasopharyngeal RT-PCR swab test for all patients, who require orthopaedic surgery (elective, emergency or non emergent trauma). We are presenting our data for the month of July 2020 in Bangalore, India. This was the time when the covid cases were increasing. We found the positive rate to be 9.52%. All were asymptomatic cases. Similar rates have been reported in an orthopaedic speciality hospital in New York for a three week period in April 2020(1). During the same time a specialized obstetric care center in New york also presented with a positivity rate of 15.4% among patients presenting for delivery(2). The false-negative rates with RT-PCR testing is high and therefore there might be a lot of under-reporting of cases(3). Ai et al got a CT chest and RT - PCR in 1014 patients with Covid-19. Of the 1014 patients, 601 of 1014 (59%) had positive RT - PCR results and 888 of 1014 (88%) had positive chest CT scans. Therefore RT-PCR may not always be a reliable test. RT-PCR has high specificity but low only moderate sensitivity.

The antibody sero-prevalence test in New Delhi (A metro similar to Bangalore) has shown a prevalence of 29%. The sero-prevalence tests have been shown to be anywhere between 0–30% world over. Therefore there are a lot of asymptomatic cases in the general population.

All patients who present with non emergent trauma are shifted to an isolation ward. Care is provided to them with all precautions. In the first half of July the turn over time for RT-PCR was 24–48 hours. Patients had to wait for a considerable amount of time before being operated on. However the turn over time has shortened and was less than a day towards the end of July. Once the RT-PCR test turns out to be negative, the patient is taken up for surgery on the next available slot.

There are multiple concerns regarding operating on a patient with an active SARS-CoV-2 infection to undergo a non emergent surgery. Any surgery affects the cell mediated immunity, which is the primary defense mechanism against viral infection(4,5). Post operative immobilisation can cause atelectasis, which can increase the chance of pneumonia. Covid –19 is associated with a prothrombotic state which can increase the chances of thromboembolic phenomenon in patients who are already at a higher risk(6–8). Catellani et al in their study believed that early fixation of hip fractures in elderly patients helps in managing their asymptomatic covid infection in a more effective way (sitting up, prone ventilation, mobilization, improved outcomes from covid as well; according to the authors)(9). In a study from China 44% of asymptomatic patients who underwent elective surgery during the incubation period of COVID-19 required ICU care and 21% of patients died(10). Hence it is imperative to have a protocol involving universal screening before undertaking any non emergent orthopaedic surgery. However asymptomatic cases have been shown to have zero mortality rate after elective surgery(1,10).

What constitutes an emergency? - The hospital administration and consultants had multiple discussions and came to conclusion regarding what constitutes an absolute emergency (i.e - what cases must be done without RT-PCR results). If the RT-PCR test turns out positive, all personnel involved in the surgery are home quarantined for a period of 7 days as per protocol and government standard operating procedures. This happened in only one of our cases and personnel involved in the case were quarantined. None of them developed symptoms and were back to work in one week’s time.

This study gives a fair idea of the number of cases in Bangalore in the month of July 2020. The number of asymptomatic cases are almost one in 10 persons is positive without symptoms in Bangalore (July 2020). Most of these cases were trauma cases and there is probably a selection bias, as people moving about are more likely to be affected with Covid-19.

## Conclusion

With these results we would like to present our protocol in managing orthopaedic surgeries during this pandemic. With a high rate of asymptomatic patients, it is pertinent to perform universal RT-PCR in all patients. Our algorithm in managing patients has proven to be effective and can be replicated with ease to continue operating and taking care of orthopaedic patients during this pandemic.

## Data Availability

All data is available

## Informed consent

Informed consent was obtained from all patients.

